# A digital intervention to improve mental health and interpersonal resilience in young people who have experienced Technology-Assisted Sexual Abuse: a feasibility clinical trial

**DOI:** 10.1101/2025.06.27.25330408

**Authors:** Sandra Bucci, Filippo Varese, Ethel Quayle, Kim Cartwright, Amanda Larkin, Cindy Chan, Prathiba Chitsabesan, Victoria Green, William Hewins, Matthew Machin, Alice Newton, Erica Niebauer, John Norrie, Gillian Radford, Cathy Richards, Marina Sandys, Victoria Selby, Sara Shafi, Jennifer Ward, Pauline Whelan, Matthias Schwannauer

## Abstract

**Background:** A digital mental health intervention (DMHI) to improve mentalisation in young people who have experienced technology-assisted sexual abuse (TASA) may reduce the risk of re-victimisation and future harm and make them more resilient and able to manage TASA-related distress. However, evidence-based interventions for TASA are nascent.

**Method:** We determined the feasibility, acceptability and safety of a 6-week mentalisation-based DMHI for young people with TASA in a pre-registered multi-centre non-randomised clinical trial (ISRCTN43130832). Young people aged 12-18 years recruited across child and adolescent mental health services in two sites completed baseline and post-treatment assessments.

**Results:** Forty-six people were recruited; 43 were allocated to the i-Minds app; 86% completed follow-up assessments. The average participant age was 15.42 years. Most participants identified as female (69.8%), White British (95.3%), a notable percentage identified as non-binary/third gender or preferred not to disclose their gender identity (16.3%), and 20.9% reported their gender did not match their sex assigned at birth. We found signals of post-treatment improvement in TASA-related post-traumatic symptoms, resilience, internalising symptoms and reflective functioning. User feedback indicated that participants generally had a positive experience of using the app, positively impacting their knowledge/understanding of their own mental health and their motivation to address their mental health difficulties. There were no related adverse events.

**Conclusions:** It is possible to recruit and retain participants for a DMHI trial of TASA. The i-Minds app was safe, acceptable and showed promising signals of efficacy on valuable outcomes. A large-scale efficacy trial is warranted to confirm and extend findings.

**Key Practitioner Message:** *What is known?:* There are currently no evidence-based treatments to support young people exposed to technology-assisted sexual abuse.

*What is new?:*

- This co-designed, multi-centre non-randomised clinical feasibility trial provides signals of potential efficacy for a digital mental health intervention to improve clinical outcomes in vulnerable young people.
- We found signals of post-treatment improvement in TASA-related post-traumatic symptoms, resilience, internalising symptoms and reflective functioning.
- User feedback indicated that participants generally had a positive experience of using the app, positively impacting their knowledge/understanding of their own mental health and their motivation to address their mental health difficulties.

*What is significant for clinical practice?:*

- A large-scale efficacy trial is warranted to confirm and extend findings.

## Background

Childhood adversities are a major risk factor for later mental health problems. With the advent of the digital age, the internet has become a place of increased risk of abuse for young people (YP), including sexual abuse (hereafter termed technology assisted sexual abuse; TASA). Approximately 15% of YP are exposed to different forms of TASA (Wachs et al., 2021). TASA can occur through any device connected to the Internet and across multiple platforms and applications and includes offences such as online grooming, image-based sexual abuse, non-consensual sexting, sextortion, and online commercial sexual exploitation. Online grooming involves adults or older youth using online communication to lure minors into sexual relationships (Gámez-Guadix et al., 2018). Image-based sexual abuse refers to taking, making, or sharing sexual images without consent (Finkelhor et al., 2022a). Non-consensual sexting also involves the sharing sexual images without permission (Krieger, 2017). Sextortion involves threatening to release sexual images to extort money or more images (Wolak et al., 2018). Online commercial sexual exploitation involves exchanging sexual images or interactions online for money or valuables (Walsh et al., 2024).

Consequences of TASA include self-harm, depression and anxiety (Kardefelt-Winther & Maternowska, 2020). While offline and TASA often co-occur, the latter brings unique harms, including self-blame and shame (Stoilova et al., 2021). Multiple factors are involved in vulnerability to being exposed to TASA. One potential risk factor is the ability to accurately recognise and understand others’ intentions and motivations in online interactions and how our own emotions can influence our responses and interactions (Bateman & Fonagy, 2010).

YP who experience emotional sequelae following internet-based victimisation, abuse, or exploitation may find understanding the intentions and motivations of others particularly challenging. The ability to attend to and have a balanced appraisal of the mental states in ourselves and others and consequently understand self/other actions based on intentional mental states is termed ‘mentalisation’ (Bateman & Fonagy, 2010). Growing evidence shows that mentalisation abilities are central to effective emotional regulation and mental well-being. The ability to mentalise one’s own experiences and those of others allows for adequate coping with external and internal stressors, regulation of affect, and the formation of stable and safe interpersonal relationships (Fonagy & Target, 2002). While no direct association between mentalisation and TASA has been tested, difficulties in mentalising processes have been linked to greater vulnerability for a range of mental health problems that are common amongst TASA survivors, including depression, anxiety, eating disorders and shame (Malda-Castillo et al., 2019). Recent systematic reviews (Malda-Castillo et al., 2019) have highlighted Mentalisation-Based Therapy (MBT), an approach that specifically aims to improve mentalising capacity, and consequently affect regulation and psychological distress, is a promising treatment approach across a wide range of clinical presentations, including groups that have previously shown limited response to psychological therapy (e.g. adolescents who self-harm). The efficacy of MBT has been more extensively trialled in adult mental health, but a growing evidence base is emerging for the efficacy of MBT in young people. For example, in a randomised controlled trial (RCT) with adolescents who self-harm, Rossouw and Fonagy (2012) found that MBT for adolescents was more effective than TAU, with a recovery rate of 44% in comparison to 17%. In a recently published pilot RCT, a brief (12 weeks) mentalisation-based intervention was acceptable in young people in receipt of CAMHS treatment and associated with significant treatment effects across a range of relevant outcomes (e.g. anxiety, self-harm behaviours, ability to regulate distressing emotions; Griffiths, et al., 2019).

In offline interactions, contextual cues are used to enhance our understanding of interpersonal interactions. In the online environment, these signals are often more opaque or non-existent, compromising a YP’s ability to mentalise, evaluate risk and assume trust in online interactions (Bleiberg et al., 2012). YP who are distressed or have difficulties with affect regulation because of their experience of being victimised, abused, or exploited online may also be at greatest risk of developing difficulties in mentalising, increasing risk for repeated victimisation and harm (Grondin et al., 2019). Early clinical trials have shown promise in mentalisation-based approaches in both group and individual formats bringing clinical benefit across a range of clinical presentations in YP. A pilot RCT conducted by Griffiths and colleagues (Griffiths et al., 2019) found that a brief (12 weeks) mentalisation-based intervention was acceptable to YP enrolled in Child and Adolescent Mental Health Services (CAMHS) and was associated with significant treatment effects on anxiety, self-harm behaviours, and the ability to regulate distressing emotions. (Rossouw & Fonagy, 2012).

When integrated within existing care pathways for YP, a Mental Health Intervention (DMHI) has the potential to increase access to support and provide timely therapeutic input which can be accessed in a familiar format for YP when traditional forms of support are not available (Bucci et al., 2019). Although the unique nature of TASA relative to other forms of abuse has been recognised in NICE guidelines in the UK for responding to child abuse and neglect, there are no internationally recommended evidence-based interventions for improving the mental health and well-being of YP who have experienced TASA. In the UK, NICE recommended further research on the efficacy of interventions aimed at improving well-being and relationships and preventing further harm following TASA. As such, we developed and co-designed the “i-Minds” app, a theory-driven DMHI underpinned by mentalisation principles, designed for young people (12-18 years) who experienced TASA. Co-design refers to the collaborative process where researchers work together with end-users and stakeholders to design and conduct research. Details of the development of the app, our method of co-design, and our registered protocol are published elsewhere (Bucci et al., 2023). We conducted a feasibility clinical trial to understand issues to be addressed in advance of conducting a powered efficacy trial of this DMHI. Additional qualitative analyses of acceptability and app usage analytics in line with the AMUsED framework (Miller et al., 2019) are reported separately.

## Methods

### Study design, participants and ethical considerations

This pre-registered trial (ISRCTN43130832;(Bucci et al., 2023) was approved by the National Research Ethics Committee (REC) of West Scotland REC 4 (approval number 22/WS/0083) and overseen by an independent oversight committee. We obtained informed written consent prior to gathering data at the baseline interview. We conducted a non-randomised feasibility clinical trial with a 6-week intervention exposure window and baseline and post-treatment assessments. Recruitment was from eight and nine CAMHS services in Edinburgh and Greater Manchester respectively, one sexual assault referral centre (SARC) in Greater Manchester, and an NHS-commissioned e-therapy provider.

Inclusion criteria were: i) aged 12-18 years; ii) exposed to TASA and report associated distress; iii) receiving support from CAMHS, SARC or an e-therapy provider and continued to be actively supported by services for the trial duration; iv) willing to use an app about TASA; v) proficient in speaking and writing in English; vi) capacity to consent; and, for e-therapy provider users only, willing to provide their username. Exclusion criteria were: i) insufficient command of English; ii) moderate learning difficulties; iii) at risk of current or recent suicidality (assessed by referrers).

### Procedure and intervention

Prospective participants were identified and approached by staff or responded to poster advertisements in waiting rooms or via digital spaces. Informed consent was obtained from those interested. Depending on participant preference, assessments took place either: i) in person (at the participant’s home or at a local clinic); ii) over the phone; iii) via remote platform.

The i-Minds app is a multi-media, interactive app consisting of a core mentalisation module and three further modules, and 13 sub-modules, available over 6-weeks. The platform is designed to support help-seeking YP-OSA (i.e. those already under the care of services or an e-therapy provider) better mentalise in the online environment. The app is intended to be used as a stand-alone platform (i.e. unsupported by a clinician / family member/trusted other). In addition to module content, participants can access a repository of multi-media material designed to support learning and promote engagement, including: videos embedded throughout the platform, audio exercises, a diary function; links to podcasts, blog posts, other useful links; real-life stories of recovery; emergency and safeguarding contacts; relaxing/breathing/mindfulness exercises (in both audio and video format), infographics on online safety, soothing images, inspiring quotes, to name a few. Participants are required to work through the mentalisation module, which is core to the theoretical orientation of the intervention, before they can access the other three modules. This module includes a ‘What is mentalizing’ animation. Hereafter, all modules and features are available. There are no limits to how often or when a participant can use the app (i.e., no restrictions) during the 6-week intervention window. That is; we did not limit how people interact with the platform, and interaction was not conditional on a participant’s response to screening items.

In line with UK Government User Research guidelines, app development was supported by a series of participatory consultations with multiple stakeholders, enabling users to influence the app’s design and functionality. We collaborated with lived experience representatives (n=7 members with lived experience of TASA aged 18-30 years) throughout the research process, from grant preparation and study material development to ethical considerations, manual adaptation, and app content design. This included troubleshooting recruitment challenges, data interpretation, and dissemination. Thirteen online LEAG meetings were held between June 2021 and July 2023 (ranging from 1-2.5hrs). Four participatory design workshops were also held and included the following phases: i) Discovery phase: introduced the project’s technical aspects and established user preferences for the app’s format, aesthetics, personalization, and resources; ii) Alpha phase: supported usability testing of app prototypes; iii) Beta phase: allowed users to try a fully functional version of the app; and iv) Live phase: enabled the LEAG to test the trial version of the app and provide feedback on their experiences. Further specific input and guidance was provided by the parents/professionals (PPAG) and a national stakeholder advisory committee. Full details of our participatory work are reported in a specific publication (Bucci et al., *in preparation)*.

Our starting point for app content development content was adapting content from a mentalisation-based manual developed by members of our team (Griffiths et al., 2019), and for app functionality was the Actissist app (Bucci et al., 2018). Content is organised into four modules: (1) mentalisation, which participants were required to work through before other app features were accessed; (2) psychoeducation (covering a range of educational material on technology use and navigating relationships and sexual experiences); (3) emotional and mental health (covering educational material and practical exercises grounded on mentalisation-based principles applied to a wide range of mental health topics, including anxiety, low mood, self-harm, positive relationships, and resilience); and (4) trauma (focused on psychological and emotional consequences of trauma, including recognising and responding to ‘triggers’, addressing and improving feelings of guilt and shame, and reducing maladaptive avoidance). Links to other areas of the app within each core topic area maximise interactivity and the user experience. Following lived experience group feedback, topics were visually represented through a tree motif to promote engagement. Further details are reported elsewhere (Bucci et al., 2023). Screenshots of the i-Minds app can be found in Figure S1.

The app could either be downloaded onto a participant’s own smartphone (iOS and Android Operating Systems) or participants were loaned a smartphone with the app preloaded. We asked participants to return loaned handsets at the end of the study. Data network charges were paid. Following an app “onboarding” session with the researcher, participants received a daily prompt in the form of a standard app notification, designed to prompt engagement. A reminder notification was sent 3 hours later where a participant did not interact with the app at the time of the initial prompt; participants could also self-initiate use of the app. At midday after each seven-day period, an additional weekly prompt invited the user to reflect on app use in the previous week. Researchers called participants 1-week after the onboarding session to troubleshoot potential technical difficulties. At the end of the 6-week intervention window, the app stopped working and the researcher either deleted the app from the participant’s phone or asked the participant to delete it themselves. Safeguarding and risk protocols were developed in collaboration with key stakeholders during the app development process to ensure the safety of both study participants and researchers throughout the trial period. No personally identifiable data was collected within the iMinds app. Information about module completion was collected to guide the unlocking of content as participants progressed through the app. De-identified usage metrics were collected using the Matomo software platform (https://matomo.org/) to inform the analysis of how people engaged with the app.

### Assessment of outcomes

#### Feasibility and engagement outcomes

Feasibility was evaluated through recruitment and retention, which were operationalised as follows: (a) number of eligible participants consenting; (b) number of participants recruited; (c) completeness of outcome measures; (d) number lost to follow-up; (e) number of services offered the intervention; and f) the extent to which YP engaged with the i-Minds app. Engagement was defined as the proportion of participants completing the intervention, dropout rates and reason for withdrawal, platform use and engagement data (the latter reported in detail elsewhere).

#### Self-report measures

We gathered descriptive clinical, demographic, technology use and satisfaction information. To examine the potential of efficacy of the i-Minds app, self-report measures were administered at both baseline and post-treatment as follows. The Reflective Functioning Questionnaire for Youths (RFQ-Y (Fonagy et al., 2016)) measures two mentalisation dimensions: certainty about mental states (RFQ-Certainty), with low scores on this scale reflecting hypermentalising and high scores reflecting more genuine mentalising, and uncertainty about mental states (RFQ-Uncertainty), with high scores reflecting a stance characterised by an almost complete lack of knowledge or ability to consider complex ways of understanding others’ and one’s own mind (hypomentalising) and lower scores characterising genuine mentalising. The Revised Child Anxiety and Depression Scale–25 item version (RCADS-25 (Ebesutani et al., 2012)) measured emotional distress. The Child Revised Impact of Events Scale (CRIES (Perrin et al., 2005)) measured online abuse related distress (by anchoring the CRIES to the YP’s TASA experience). The Difficulties in Emotion Regulation Scale-Short Form (DERS-SF (Kaufman et al., 2016)) measured emotional regulation. The Interpersonal Sensitivity Measure (ISM (Boyce & Parker, 1989)) measured interpersonal sensitivity. The Connor-Davidson Resilience Scale-10 item version (CD-RISC-10 (Campbell-Sills & Stein, 2007)) measured resilience. At baseline, we also administered the Problematic and Risky Internet Use Screening Scale (PRIUSS;(Jelenchick et al., 2014)), (an 18-item scale scored on a 5-point Likert scale (1=never; 5=very often) to measure problematic internet use, and the Experiences in Close Relationships Scale–Revised Child version (ECR-RC (Marci et al., 2019)) to measure the views/attitudes towards close interpersonal relationships.

At follow-up, we gathered further app feedback using the user version of the Mobile Adherence Rating Scale (uMARS (Stoyanov et al., 2016)), a 20-item multidimensional objective measure of app usability, with four subscales: engagement, functionality, aesthetics, information quality.

Demographic, clinical, and CRIES were administered with the support of the research worker; all other measures were administered either with support of the research worker or self-directed. Assessors received training and ongoing supervision in all study procedures. Participants were remunerated £20 per study assessment.

To measure safety of the intervention, adverse events (AEs) were monitored and reported using standardised operating procedures in line with UK Health Research Authority and local research and development policies of participating NHS organisations. All AEs were assessed for relatedness to the intervention and/or trial procedures by a senior clinical researcher and were monitored by an independent Data Monitoring and Ethics Committee.

#### Analytic approach

Data were entered and managed on the REDCap platform (Harris et al., 2009). In line with the CONSORT statement for pilot and feasibility trials, recruitment and retention data was summarised using descriptive statistics (Eldridge et al., 2016). Descriptive statistics were also used to summarise the completeness of baseline and follow-up assessment instruments (i.e., % of missing data across partially completed measures and whole questionnaires), and the number and nature of AEs observed over the course of the trial. As per protocol, the trial was not powered to detect significant post-intervention changes at follow-up; hence no inferential test was conducted. Summary statistics (Mean, SD, median, range) were tabulated for each measure at each time point (baseline, post-treatment). Pre- and post-intervention questionnaire data were further examined by computing change scores and associated 95% confidence intervals (CIs), and further analysed at the individual participant level using the Leeds Reliable Change Indicator (RCI; (Morley & Dowzer, 2014) to determine the number of participants who reliably improved on clinical measures. Deprivation index scores estimated for each participant based on their postcode of residence were calculated by standardising the deprivation scores between the England and Scotland based scoring range using z transformation. The standardised scores were converted into deciles by ranking the relative Scottish and English data zones into 10 equal groups. As we aimed to evaluate measure completeness, no attempt was made in impute missing data, and all descriptive analyses refer to observed data only. Free-text data for uMars items were analysed using an inductive content analysis approach (Elo & Kyngäs, 2008).

## Results

### Participant characteristics

Demographic characteristics of the sample are reported in Table S1. Participants average age was 15.42 years. most participants (69.8%) identified as female and White British (95.3%). A notable percentage identified as non-binary/third gender or preferred not to disclose their gender identity (16.3%) and 20.9% reported their gender did not match their sex assigned at birth. The Scottish and English Index of Multiple Deprivation is the Governments’ respective standard approach to identify areas of multiple deprivation.

Data zones for Scotland are ranked from 1 (most deprived) to 6,976 (least deprived), and for England are ranked 1 (most deprived area) to 32,844 (least deprived area). Participants were evenly distributed across deprivation index centiles (see Table S2 and Figure S2). As illustrated in Figures S3 and S4, a sizable proportion of participants continued to have ongoing exposure to potentially harmful online materials and interactions in the year prior to their participation in the i-Minds trial.

### Recruitment and retention

The CONSORT diagram is shown in Figure S5. Recruitment and trial enrolment took place over 10 months between May 2022 and March 2023. In this period, 147 YP were screened for eligibility by staff within participating clinical services; 72 referrals were made. The conversion rate from screening to referral was significantly superior for participants screened by NHS services compared to the e-therapy provider (94.5% of NHS prospective participants were referred to the trial, vs 4.1% for the e-therapy provider). We approached 24 services across Manchester (15) and Edinburgh (9); one service declined to support the study due to staffing pressures. Of the 24 services who agreed to support the study, 17 services (Manchester = 9; Edinburgh = 8) referred at least one young person to the study. 65 participants were deemed eligible for the trial; 46 consented to trial participation. Three participants withdrew consent prior to baseline completion (one due to concerns related to burden of study assessments; two due to busy schedules). 43 YP completed baseline assessment and were allocated to the i-Minds intervention. Site differences were evident in terms of recruitment performance; however, these were primarily due to staff vacancies effecting one site.

Overall, 86% of recruited participants completed the follow-up assessment (six were lost to follow-up). A breakdown of retention rates by site is reported in Table S3 indicating higher retention at the Manchester (92.3%) relative to the Edinburgh (82.1%) site. While we planned to examine potential differences in retention across key demographic groups (e.g., ethnicity, gender), these analyses were not performed as they would not lead to reliable results in this demographically homogeneous sample.

### App usage

Forty-one YP were onboarded. Number of accessed and completed modules were as follows: 95% users (39/41) completed the mandatory *mentalisation* module; 39% (16/41) users completed, 44% (18/41) accessed, and 17% (7/41) never accessed the *psychoeducation* module; 39% (16/41) users completed, 34% (14/41) accessed, and 27% (11/41) never accessed the *emotional and mental health module*; 46% (19/41) users completed, 32% (13/41) accessed, and 22% (9/41) never accessed the *trauma* module. Most participants (91%) accessed the app via their own phone; 9% of the sample requested a study handset (see Table S4).

### App acceptability and user satisfaction

Despite variation in feedback, participants (n=37) generally reported a positive experience using the app (see Table S5). Participants rated the app as not excessively burdensome (e.g., in terms of required work and time, see items 1-2; potential emotional burden/stress that may be caused by the sensitive topics addressed by the app, see items 6 and 9). Participants generally rated their experience of using the app as enjoyable (item 8), that it made them ‘feel better’ (item 10) and that other YP would find the app easy to use (item 5). 64% participants reported that they would use the app for longer than the 6-weeks intervention window used in the current trial.

Acceptability ratings from the uMARS measure (see Table S6; n = 36) shows that the app content was rated as highly relevant and appropriate for YP; however, other items under the ‘Engagement’ domain suggest that further improvement may be needed to enhance user engagement, particularly in relation to the extent the app could be customised and made more entertaining to use. The other uMARS domains attracted consistently high scores in relation to the functionality and aesthetics of the app, as well as the quality, quantity and credibility of the information/content covered. Further subjective uMARS feedback (see Figure S6) indicated that 17% of users gave the app a ‘5 stars’ mark (the best app I have ever used), 58% ‘4 stars’ and 22% ‘3 stars’ (average). Most participants reported that they would recommend the app to YP with similar experiences, and that they would continue to use the i-Minds app in the following 12 months if TASA was relevant to them. Most participants indicated that they did not share/talk about the app content with family members/parents/carers or professionals involved in their care. Content analysis of uMARS open feedback showing what users liked and suggestions for improvement to the app, mapped according to the uMARS domains, is shown in Table S7.

Data on the perceived impact of the i-Minds app on users’ attitudes towards and understanding of their mental health difficulties is summarised in Table S8 and Figures S7 and S8. Using the app was rated by a considerable proportion of participants as positively impacting their knowledge / understanding of their own mental health, their motivation to address their mental health difficulties, and seeking future help for such difficulties.

However, feedback on whether the app impacted more broadly on users’ awareness of the importance of their mental health, their attitudes toward improving mental health problems, and actively engaging in activities that could help their mental health, was mixed.

### Change on self-report clinical measures

As illustrated in Table S9, many participants presented clinically significant difficulties across several domains, including elevated post-traumatic symptoms linked to their TASA experiences (CRIES total scores > 20; Mean = 44.78, SD = 13.01) and problematic internet use (PRIUSS total scores > 25; Mean = 48.19, SD = 15.97). Due to high levels of diversity in terms of gender identity in this sample, RCADS-25 total scores are presented as opposed to *T* scores for the relevant gender and age groups; the RCADS-25 scores therefore cannot be interpreted clinically but nonetheless are suggestive of elevated internalising problems in this sample (i.e., elevated anxiety and depression; Mean = 38.71, SD = 16.02).

Regarding measure completeness, the percentage of missing ranged between 0% to 14% at baseline, and between 16% to 21% at follow-up. Missing data were more pronounced when measure completion was self-directed vs supported by a research worker. Change scores for the RFQ subscales (and their associated 95% CIs) suggested that the RFQ-Certainty subscale scores remained relatively unchanged between baseline (Mean = 1.74, SD = 2.52) and follow-up (Mean = 1.97, SD = 2.81), but there was some evidence of improved mentalisation ability at follow-up on the RFQ-Uncertainty subscale (as indicated by the 95% CI narrowly including the point of null effect; Mean = −1.12, SD = 3.76; 95% CI = −2.43,0.19). This trend was confirmed by RCI analyses (Table S10) indicating that, while 85% of participants did not present any reliable change on RFQ-Certainty, reliable changes were evident for a larger number of participants on RFQ-Uncertainty (29% of the sample demonstrating a reliable improvement, but also 12% of the sample displaying a deterioration on this measure).

Furthermore, we observed a moderate correlation between RFQ-Certainty and RFQ-Uncertainty of *r*= −0.47, suggesting that increases in Certainty are associated with decreases in Uncertainty and that Uncertainty is more dynamic (i.e., there is more stability in certainty scores and more change in uncertainty scores).

Further potential improvement at follow-up were also observed for the CRIES total score and intrusions subscale of the CRIES (suggesting improved TASA-related post-traumatic symptoms, with 19% participants achieving a reliable improvement in the CRIES total score RCI analyses, and 8% a deterioration in scores) and the CD-RISC-10 (suggestive of amelioration in resilience at follow-up, with 18% participants achieving a reliable improvement on this measure). Further signals of benefit were observed for the RCADS-25 scores (total and the major depression scores). However, these potential changes could not be examined further via RCI analyses due to issues in calculating age- and gender-adjusted *T* scores for this sample (which prevents comparisons between the current data and RCADS-25 comparative data from the published literature, as required for the calculation of RCIs). There was no indication of change from baseline on the remaining self-report measures administered at follow-up (DERS-SF and ISM).

### Safety

We recorded 17AEs, all unrelated to trial procedures and the intervention. Two of these unrelated events were rated as SAEs (physical health hospitalisation; A&E visit for intentional overdose).

## Discussion

i-Minds is a new DMHI for YP with distress associated with TASA. At two UK sites, we successfully identified and recruited eligible participants, with high levels (>80%) of retention at follow-up. A larger scale evaluation of the i-Minds intervention appears feasible and warranted. The levels of app engagement and generally positive user feedback suggests that i-Minds is not only tolerable but also a valued and relevant support tool. Given the well-noted problem of sustained app use over time (Torous et al., 2018) perhaps due to misalignment between researcher’s aims and the end-user’s needs (Byambasuren et al., 2018), our process of co-design might explain the high levels of satisfaction with the app and study procedures.

While our White British sample reflects the gender and ethnicity of our target population in the reported literature (International Watch Foundation, 2023), Business Intelligence data (the collection, analysis, and presentation of healthcare data to support decision-making and improve healthcare services) from NHS Mental Health services in our two recruitment sites showed that around 80% of YP registered with CAMHS are also White British. This suggests that our trial participants were representative of the clinical services from which we recruited, though not necessarily of the broader population. The gender difference observed in our sample may reflect lower levels of disclosure among males and the lower reported incidence of TASA in this group. Additionally, research suggests that YP from ethnic minority backgrounds often face unique barriers to accessing mental health services, leading to their under-representation in clinical research (Finkelhor et al., 2022b). While current prevalence rates indicate that females are at greater risk of and more vulnerable to TASA, some reports show that YP from ethnic minority groups and of different sexual orientations face an even greater risk (Thorn, 2021). Despite this finding, we observed a higher-than-expected proportion of participants identifying as non-binary, third gender, or as a gender that did not match their sex assigned at birth. This aligns with data suggesting that LGBTQ+ and sexual minority YP may be at greater risk of TASA. Future research should further explore these diversity aspects, particularly extending to sexual orientation, which was not assessed in the current study. Additionally, efforts should be made to improve the inclusion of ethnically diverse populations in TASA-related studies to ensure findings are more representative and that interventions are culturally responsive.

Data from our rigorous reporting of AEs suggest that the i-Minds app has a promising safety profile in this vulnerable young group, as no SAEs were related to the intervention or trial procedures. Findings sit alongside the YP’s literature that digital approaches to supporting YP’s mental health are feasible, safe and acceptable (Hollis et al., 2017).

The findings also support the potential for efficacy of the i-Minds app. Our findings indicated there were promising, although modest, pre-post changes in self-report instruments assessing post-traumatic symptoms linked to TASA, resilience, mood, and the reflective functioning domain uncertainty. Self-report data also indicated that for a considerable proportion of participants, using the app was perceived as having a positive impact on their knowledge / understanding and motivation to address their mental health difficulties, and inclination to seek future support for such difficulties. As the app content was underpinned by mentalisation-based principles and content, the app might exert its potential effect on improving YP’s mentalisation skills. Alternatively, as highlighted by our lived experience group, it is also possible that the effect might be driven by the non-shaming, destigmatising stance reflected in the app content, as well as the more generic psychoeducational content of the intervention, which may help YP to engage more readily and openly with difficult TASA experiences. Mechanistic research may be integrated into a future efficacy trial to clarify how i-Minds may bring benefit on valued outcomes for the target population and inform subsequent measures to maximise the effectiveness of this treatment approach for YP distressed by TASA. A future trial might also consider extending the treatment window to determine whether, in line with feedback received in this initial evaluation, user engagement continues beyond 6-weeks and whether ongoing use is associated with sustained treatment gains. As YP in this study were registered with mental health services, this might also account for the observed change on clinical measures.

### Limitations

Some considerations must be noted. As there was no randomisation and active/inactive control arm, we cannot establish whether any apparent gain is attributable to using the i-Minds app. The signals of efficacy observed across varied outcomes are nonetheless encouraging, particularly considering the high levels of ongoing exposure to potentially harmful online materials and interactions reported by study participants at baseline, which may mask or diminish possible treatment gains of an intervention specifically designed to target the *sequelae* of such online arms. Future evaluations could benefit from using recently proposed trial design and analysis frameworks that account for events occurring after treatment begins, or ‘intercurrent events’ (such as ongoing online harms or victimization), that may affect treatment efficacy. One such framework is the estimands paradigm from causal inference literature, as introduced in the ICH E9 (R1) Addendum on Estimands and Sensitivity Analysis in Clinical Trials (Clark et al., 2022), which provides a structured approach to defining and analysing treatment effects while considering these events. While future evaluations of the i-Minds app should involve a RCT design, we note that there is no standard evidence-based intervention or treatment pathway for TASA and associated distress, which might pose challenges for the selection of an appropriate comparator to test efficacy.

Findings are limited to largely White British help-seeking females. Those who referred were filtered by clinicians, which may introduce bias into the sample as clinicians made decisions about referrals based on views such as whether a YP was too distressed or not ‘ready’, or whether the DMHI ‘fit’ with the treatment model they were using; this possibly explains the observed lack of diversity (e.g., ethnicity, gender) observed in the current sample. That is, research indicates that clinicians may hesitate to discuss topics such as sexual orientation and sexual experiences when children and their family members are from certain demographic groups due to concerns about cultural sensitivities and perceived taboos (Braybrook et al., 2023). As NHS services do not have a clinical pathway for TASA (Schmidt et al., 2023), findings do not necessarily reflect all YP who experience TASA.

## Conclusion

i-Minds is a feasible, acceptable and safe DMHI for TASA. Future evaluations should consider additional outcomes valued by the target group and identify a plausible mechanistic target for the i-Minds DMHI. While we focused recruitment on secondary/tertiary-level services, the trial revealed the need for a diversified recruitment approach to reach vulnerable young people supported by social care or the educational sector, who might not access mental health services routinely. Further work is needed to differentiate the forms of TASA and their consequences, enabling young people to better navigate and personalize the i-Minds app content. This will also help services understand whether a DMHI like i-Minds should complement or stand apart from existing trauma-informed care models. This trial is a significant step toward developing both an evidence base and a digital platform to support young people exposed to TASA, demonstrating how clinical feasibility trials can underpin digital healthcare with theoretical frameworks and lived experience involvement.

## Data Availability

The data set generated in this study is not publicly available owing to ethical restrictions for personal health information; however, limited, deidentified data may be made available from the corresponding author on reasonable request.

## Acknowledgements

The authors thank the clinicians who referred to and supported the delivery of the study, and the young people who took part in this trial. We are grateful for the support of our Trial Steering Committee and Data Monitoring and Ethics Committee. We also thank the members of the Lived Experience Advisory Group, the Parents and Professionals Advisor Group and the National Stakeholder Advisory Committee for their input, advice and guidance.

## Author contributions

Conceptualization: Bucci, Varese, Quayle, Schwannauer. Formal analysis: Norrie.

Funding acquisition: Bucci, Varese, Quayle, Schwannauer, Cartwright, Chitsabesan, Green, Machin, Richards, Whelan, Norrie.

Investigation: Chan, Hewins, Niebauer, Sandys, Ward, Cartwright, Larkin. Methodology: Norrie, Bucci, Varese, Schwannauer, Quayle.

Project administration: Cartwright, Larkin. Resources: Not applicable.

Supervision: Bucci, Varese, Quayle, Schwannauer, Richards, Radford, Shafi, Chitsabesan, Larkin, Cartwright.

Writing – original draft: Bucci, Varese.

Writing – review and editing: All authors critically revised the manuscript for intellectual content and approved the final version of this paper.

Other contributions – PPIE: Green, Newton, Whelan, Cartwright, Quayle, Larkin, Bucci.

Other contributions – Intervention development: Selby, Bucci, Schwannauer, Varese, Quayle. Other contributions – Software development: Machin, Whelan.

The views expressed in this paper are those of the authors and may not reflect those of their organisational affiliations, nor of other collaborating individuals or organisations.

## Ethical considerations

The relevant Institutional and Health Research Authority ethics approvals were granted (REC Number: 21/WS/0160) and the protocol was registered at ClinicalTrials.gov (ClinicalTrials.gov.co.uk). Identifier: NCT05006053). All procedures performed in this study involving human participants were in accordance with the ethical standards of the institutional and/or national research committee and with the 1964 Helsinki Declaration and its later amendments or comparable ethical standards. Written informed consent to participate in this study was obtained from all participants. The procedures followed were in accordance with the English and Scottish legislation of the UK Health Regulation Authority.

---

**Figure S1.**
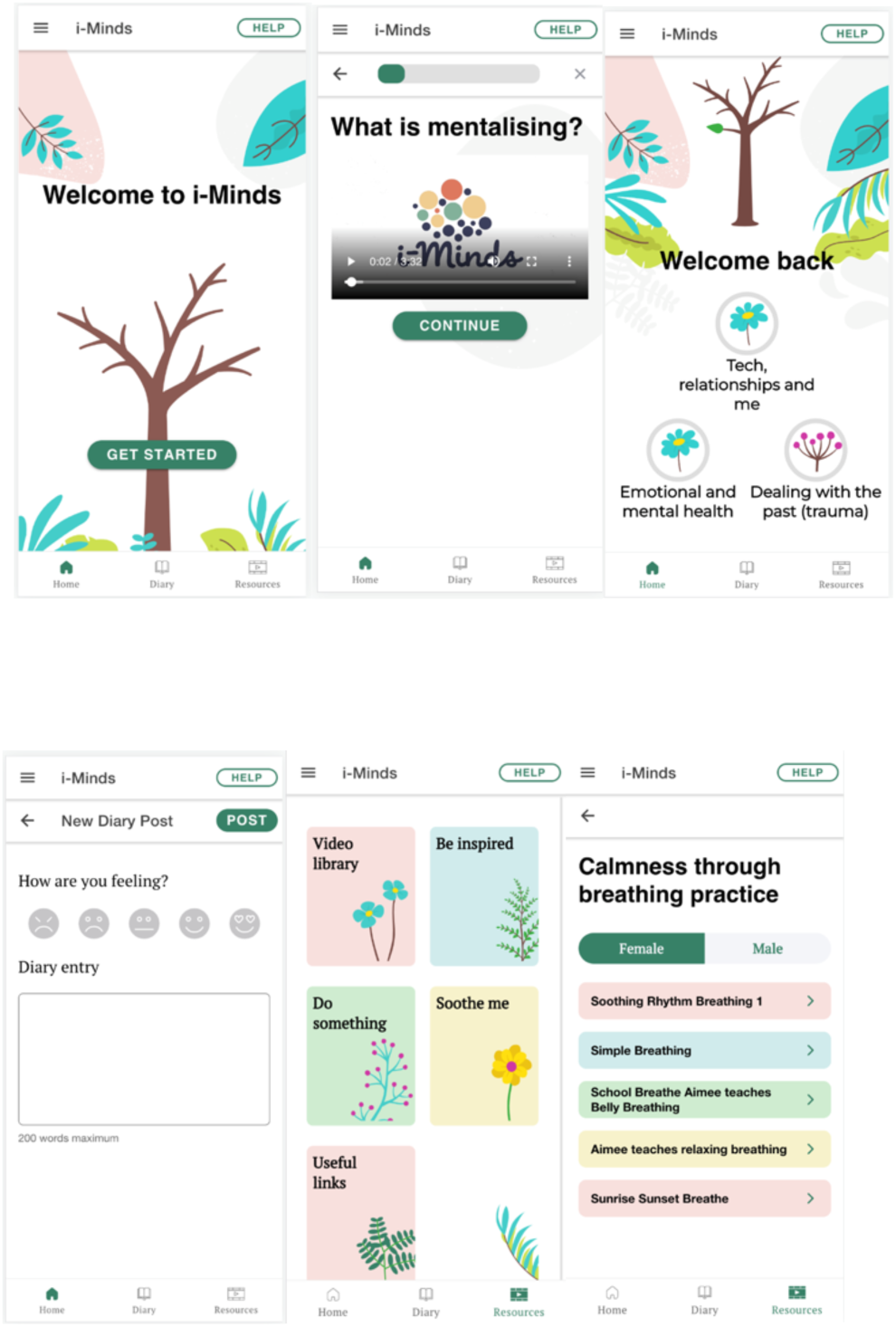
Screenshots of the i-Minds app

**Figure S2.**
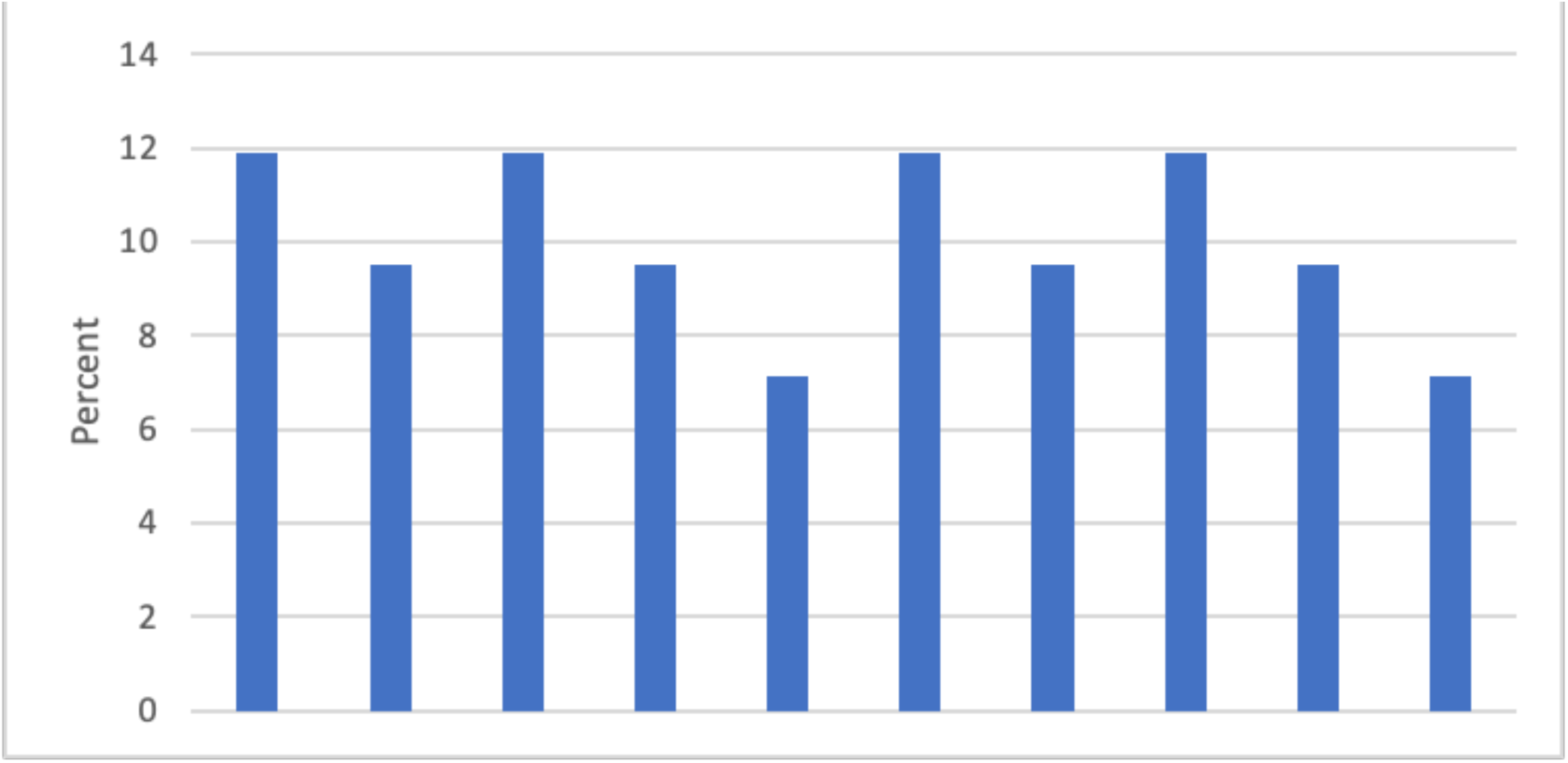
Standardised deprivation index scores

**Figure S3.**
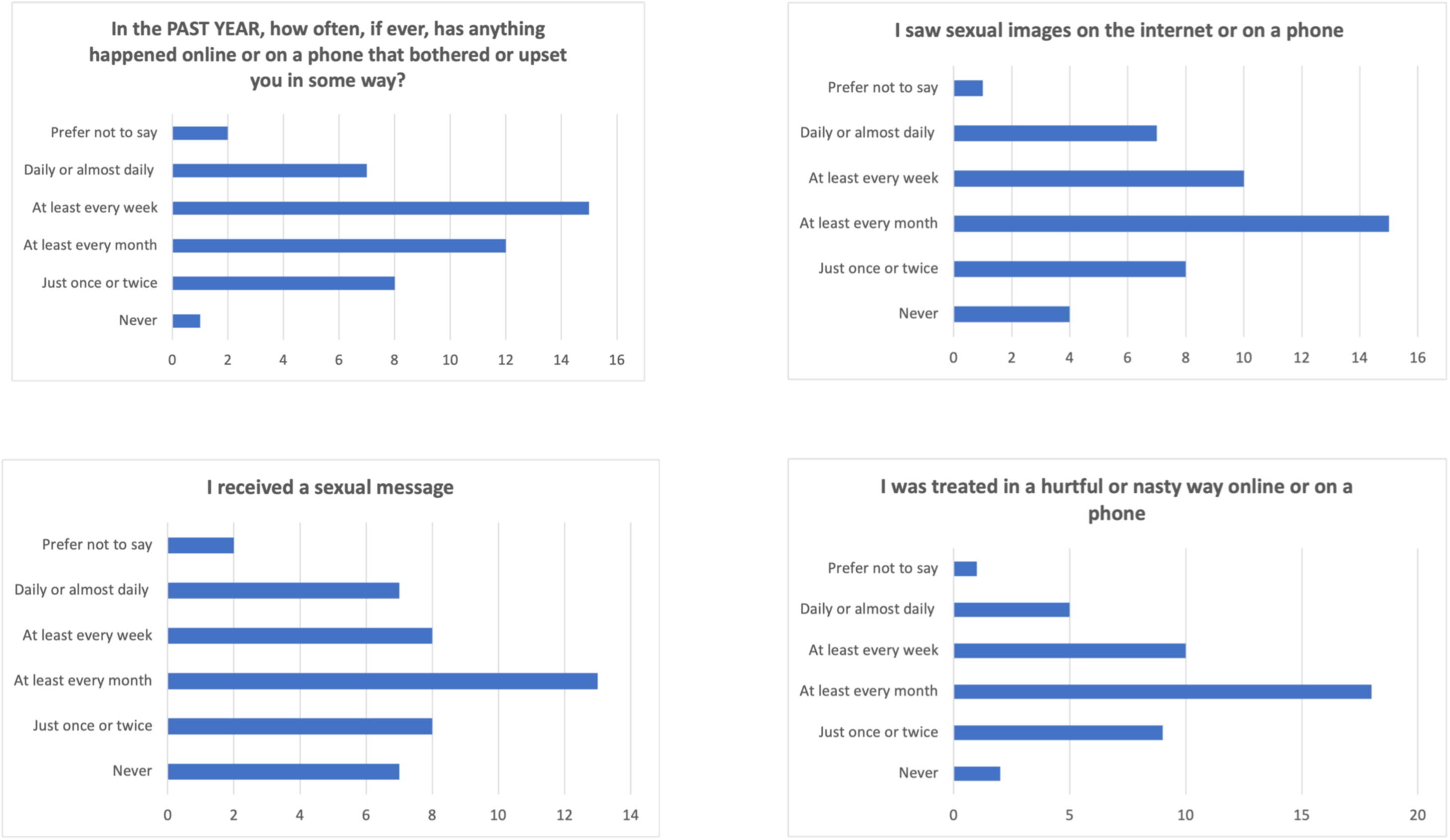
Bar charts illustrating self-reported frequency of exposure to potentially harmful online materials and interactions in the previous year (Items 1-4)

**Figure S4.**
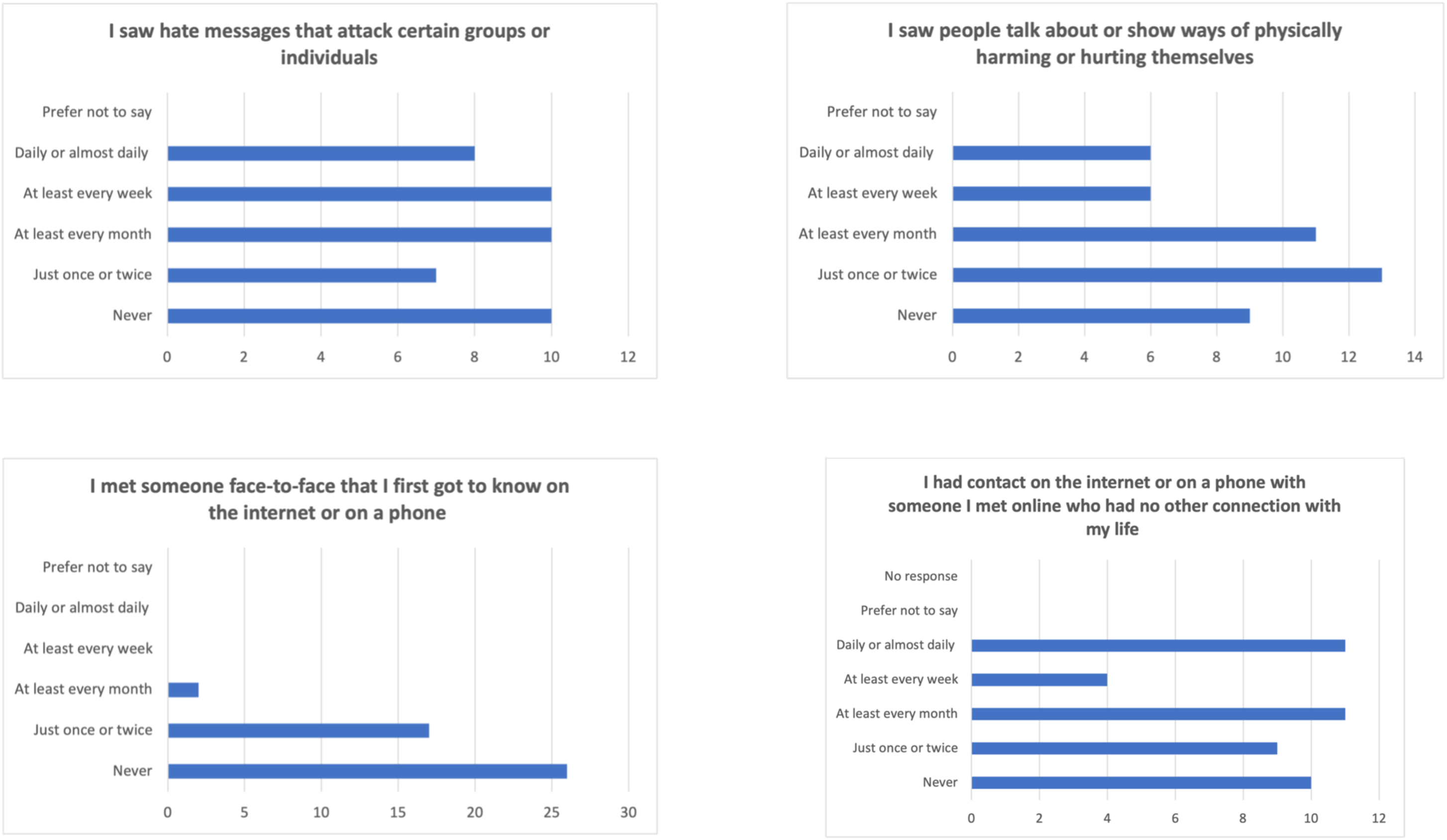
Bar charts illustrating self-reported frequency of exposure to potentially harmful online materials and interactions in the previous year (Items 5-8)

**Figure S5.**
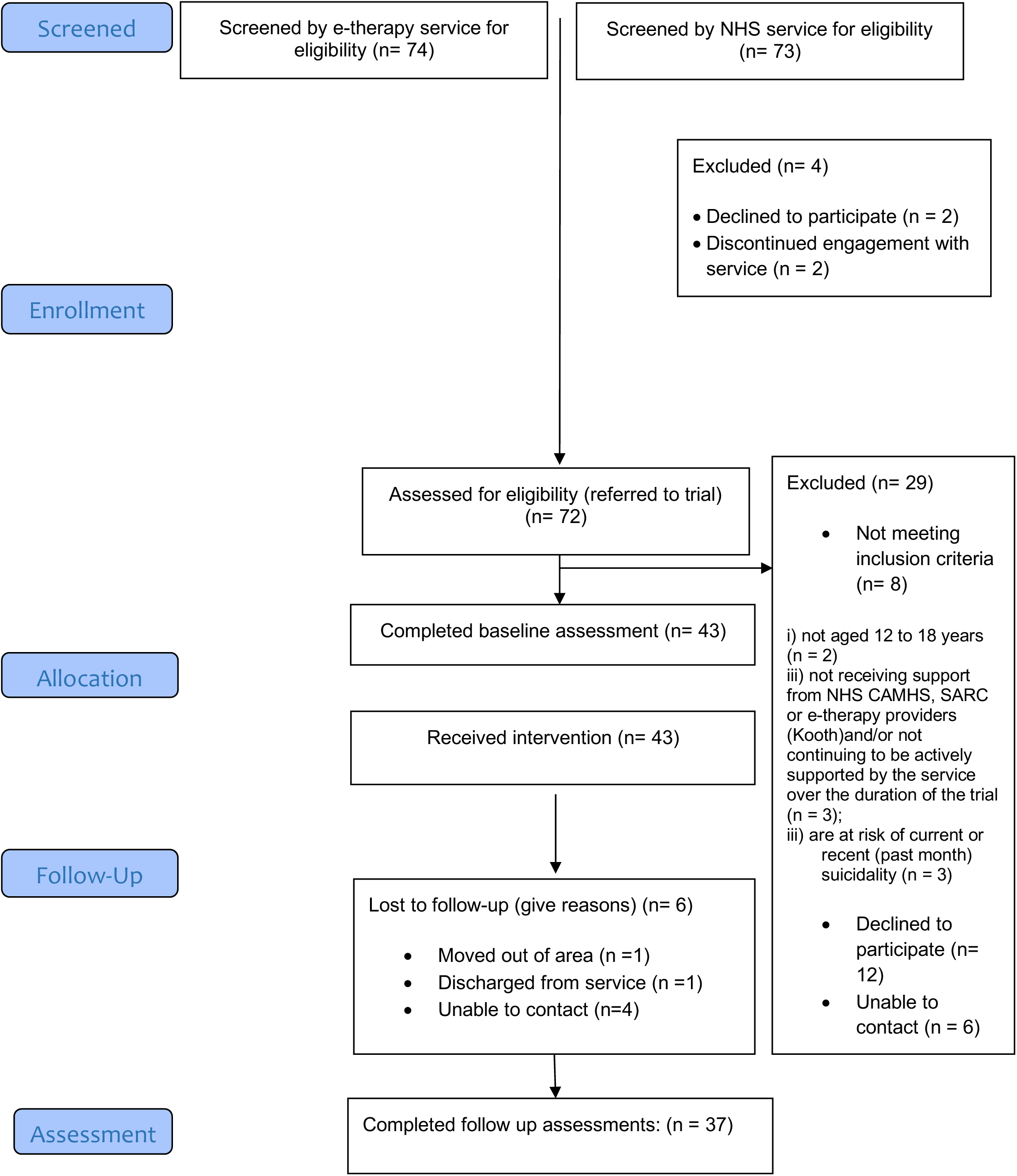
Consort diagram illustrating participant flow through the i-Minds trial

**Figure S6.**
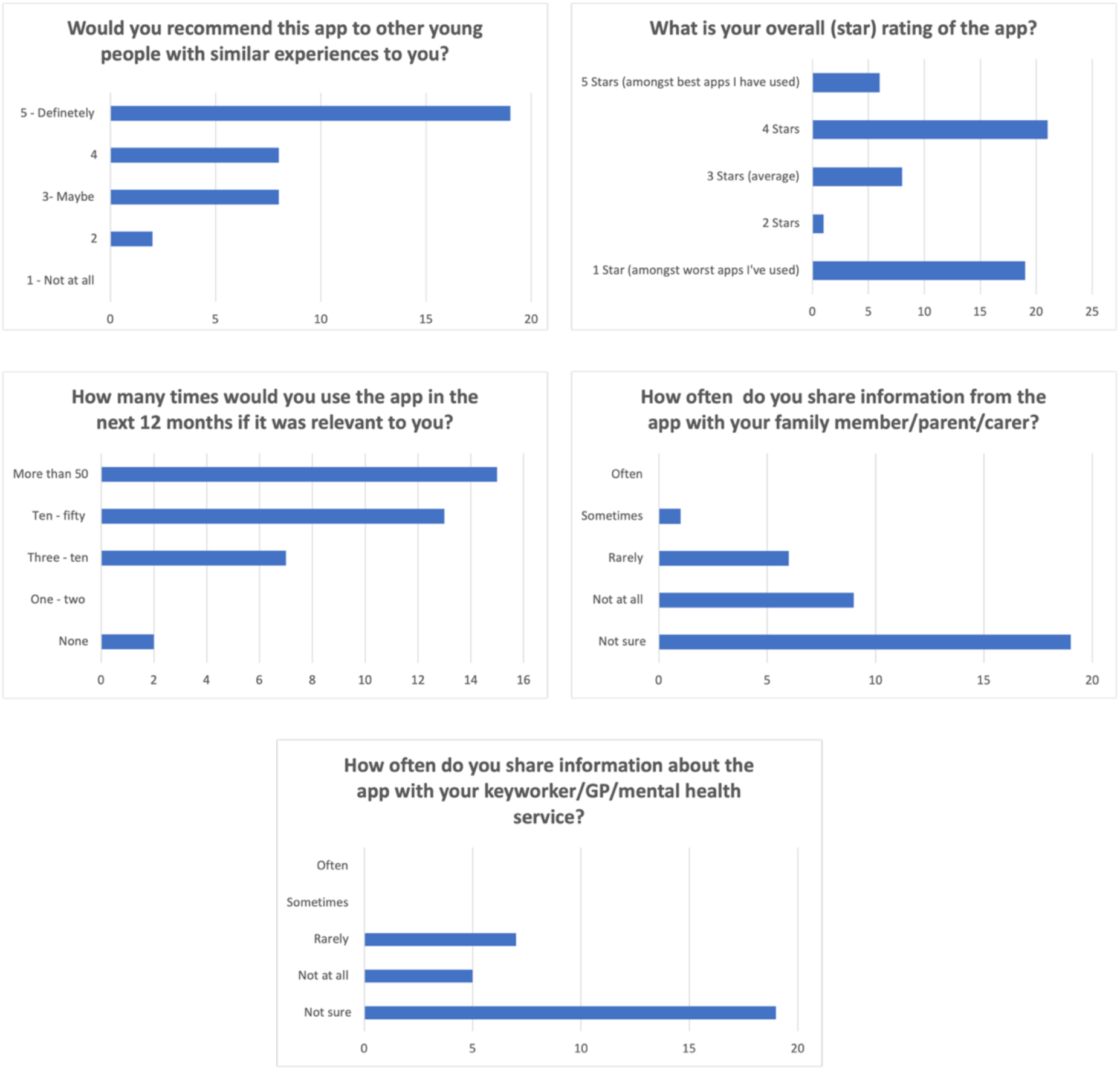
Bar charts illustrating the additional subjective feedback provided as part of the uMARS

**Figure S7.**
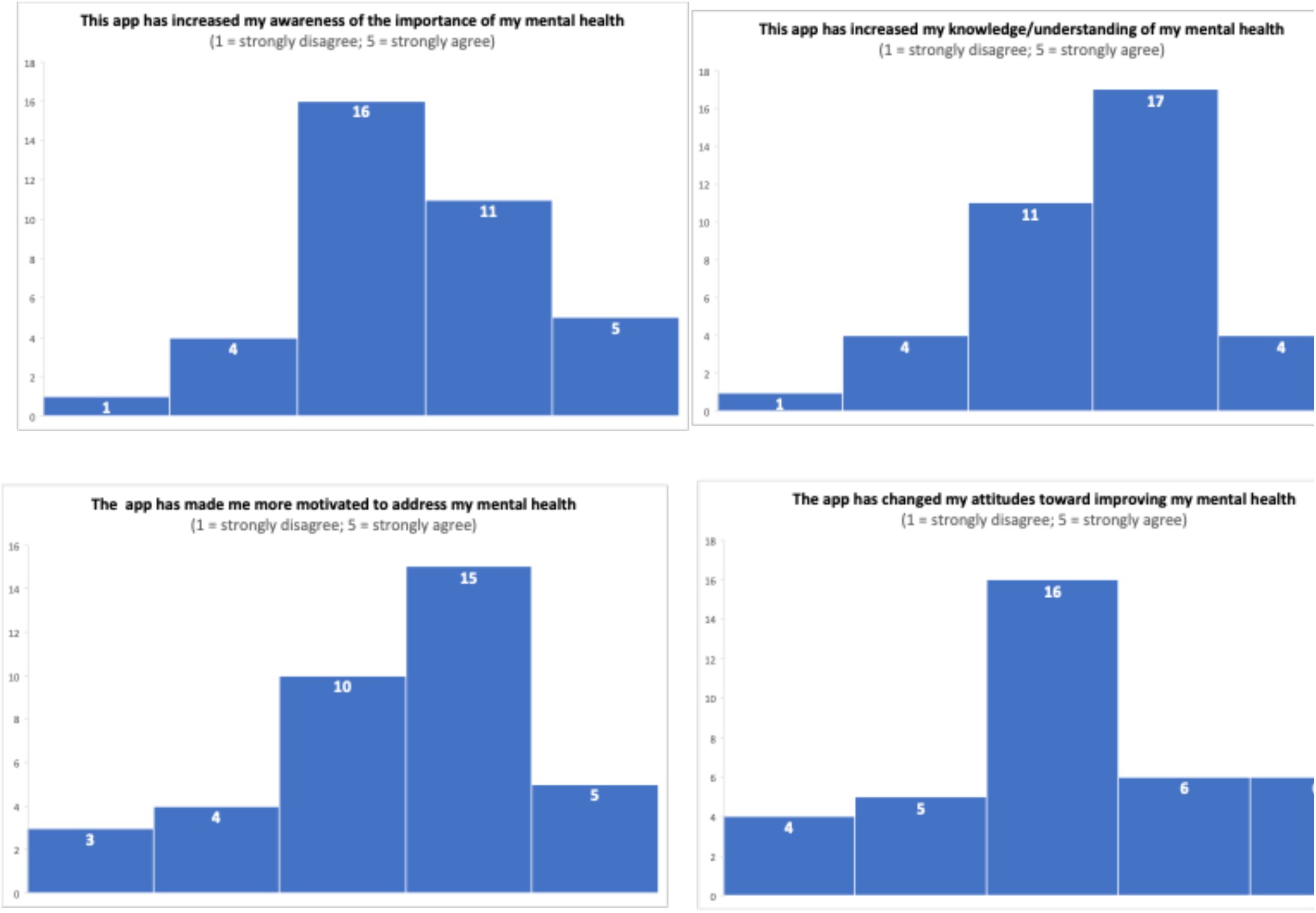
Histograms for impact of i-Minds app on mental health (Items 1-4)

**Figure S8.**
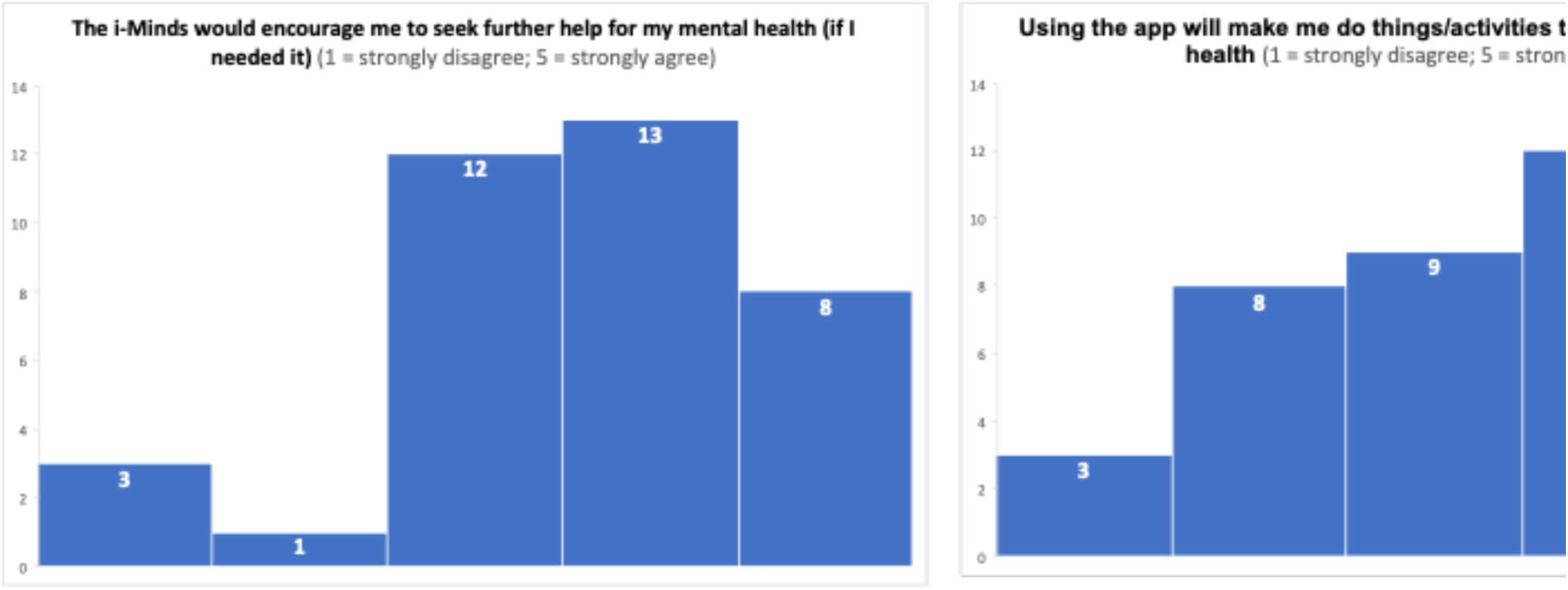
Histograms for impact of the i-Minds app on mental health (Items 5 and 6)

**Table S1.** Baseline Characteristics and service use.

| Characteristic |  | N (%) |
| --- | --- | --- |
| Age (years) | Mean (min-max) | 15.42 (12.03 – 18.06) |
| Gender | Man / Male (n/%) | 6 (13.9%) |
|  | Woman / Female (n/%) | 30 (69.8%) |
|  | Non-binary / Third gender | 4 (9.3%) |
|  | I would rather not say / unsure | 3 (7.0%) |
| Gender matches sex assigned at birth | Yes | 33 (76.7%) |
|  | No | 9 (20.9%) |
|  | Prefer not to say | 1 (2.3%) |
| Ethnicity | White British | 43 (95.3%) |
|  | Asian/Asian British | 1 (2.3%) |
|  | Black British | 1 (2.3%) |
| Deprivation Index | Range | Scotland: 278 – 6,933<br>England: 4,282 – 32,503 |
| Baseline service use and use of internet / being online |  | N (%) |
| Currently supported by CAMHS | No | 2 (4.4%) |
|  | CAMHS / Child & Adolescent Mental Health Team | 34 (75.6%) |
|  | Sexual Assault Referral Centre (SARC) | 3 (6.7%) |
|  | e-therapy provider | 3 (6.7%) |
| Length of time<br>receiving help from<br>services (months) | [N] Mean (SD) min-max | 16.09 (22.89) 1 - 96 |

**Table S2.** Standardised deprivation index scores.

| Decile | Frequency | % |
| --- | --- | --- |
| 1 | 5 | 11.6 |
| 2 | 4 | 9.3 |
| 3 | 5 | 11.6 |
| 4 | 4 | 9.3 |
| 5 | 3 | 7.0 |
| 6 | 5 | 11.6 |
| 7 | 4 | 9.3 |
| 8 | 5 | 11.6 |
| 9 | 4 | 9.3 |
| 10 | 3 | 7.0 |
| Total | 42 | 97.7 |

\*Each decile represents approximately 10% of England/Scotland's population from the most deprived 10% of the population to the least deprived 10%. Data zones in decile1 fall within the most deprived 10% of areas nationally, and data zones in decile 10 fall within the least deprived 10% of zones nationally.

**Table S3.** Breakdown of retention by recruitment site.

| **Site** | **Baseline**<br>**completed** | **Follow-up**<br>**completed** | **% retention** |
| --- | --- | --- | --- |
| **Edinburgh** | 28 | 23 | 82.1% |
| **Manchester** | 13 | 12 | 92.3% |
| **e-therapy provider** | 2 | 2 | 100.0% |
| **Total** | 43 | 37 | 86.0% |

**Table S4.** Intervention accessed via own phone or study handset.

|  | **Edinburgh** | **Manchester** | **e-therapy**<br>**provider** | **Total** |
| --- | --- | --- | --- | --- |
| **Own Phone** | 25 | 12 | 2 | 39 |
| **Study Handset** | 3 | 1 | 0 | 4 |
| **Handset returned post-**<br>**intervention** | 2 | 1 | 0 | 3 |
| **Handset not returned**<br>**post-intervention (reason)** | 1 (taken by police as<br>evidence in an<br>ongoing investigation) | 0 | 0 | 1 |

**Table S5.** Descriptive statistics for the general app satisfaction items.

| Item* | N | Mean<br>(SD) | Min-Max | Median |
| --- | --- | --- | --- | --- |
| 1. Did using the app take a lot of work? | 37 | 2.16 (1.57) | 1-7 | 2 |
| 2. Did using the app take up a lot of time? | 37 | 2.49 (1.30) | 1-7 | 3 |
| 3. Was it difficult to keep the/your Smartphone with you or carry it around? | 37 | 1.32 (1.27) | 1-7 | 1 |
| 4. Did you ever lose or forget the Smartphone? | 37 | 1.81 (1.76) | 1-6 | 1 |
| 5. Do you think other young people would find the app easy to use? | 37 | 5.78 (1.47) | 2-7 | 6 |
| 6. Overall, was using the i-Minds app Stressful | 37 | 2.03 (1.66) | 1-6 | 1 |
| 7. Overall, was using the i-Minds app challenging | 37 | 2.24 (1.59) | 1-7 | 2 |
| 8. Overall, was using the i-Minds app enjoyable? | 37 | 4.73 (1.54) | 1-7 | 5 |
| 9. Did using the i-Minds app make you feel worse? | 37 | 1.89 (1.35) | 1-6 | 1 |
| 10. Did using the i-Minds app make you feel better? | 37 | 4.59 (1.72) | 1-7 | 5 |
| 11. Do you think it would be helpful for your keyworker/GP to see how you answered the questions in the app? | 37 | 3.38 (1.98) | 1-7 | 4 |

*\*All items rated on a 7-point Likert scale (1 = Not at all; 7 = Very much so)*

**Table S6.** Descriptive statistics for the uMARS.

|  | N | Mean (SD) | Mi<br>n. | Max. | Median | Range of<br>possible<br>scores |
| --- | --- | --- | --- | --- | --- | --- |
| Subscale / Item |  |  |  |  |  |  |
| Engagement | 2 | 17.06 (3.62) |  | 23 | 17.50 | 5-25 |
| 1. Entertainment | 36 | 3.33 (0.82) | 1 | 5 | 3 |  |
| 2. Interest | 36 | 3.89 (0.95) |  | 5 | 4 |  |
| 3. Customisation | 36 | 2.35 (1.31) | 1 | 5 | 2 |  |
| 4. Interactivity | 35 | 3.58 (1.34) | 1 | 5 | 4 |  |
| 5. Target Group | 36 | 4.39 (0.72) | 3 | 5 | 5 |  |
| Functionality | 36 | 17.78 (1.90) | 12 | 20 | 18 | 4-20 |
| 6. Performance | 36 | 4.46 (0.80) | 2 | 5 | 5 |  |
| 7. Ease of use | 36 | 4.43 (0.76) | 2 | 5 | 5 |  |
| 8. Navigation | 36 | 4.33 (0.63) | 3 | 5 | 4 |  |
| 9. Gestural Design | 36 | 4.57 (0.65) | 3 | 5 | 5 |  |
| Aesthetics | 36 | 12.83 (1.48) | 9 | 15 | 13 | 3-15 |
| 10. Layout | 36 | 4.54 (0.65) | 3 | 5 | 5 |  |
| 11. Graphics | 36 | 4.16 (0.69) | 3 | 5 | 4 |  |
| 12. Visual Appeal | 36 | 4.08 (0.83) | 2 | 5 | 4 |  |
| Information | 36 | 15.97 (2.86) | 6 | 20 | 16.50 | 4-20 |
| 13. Quality of Information | 36 | 4.51 (0.77) | 2 | 5 | 5 |  |
| 14. Quantity of<br>Information | 36 | 4.19 (1.10) | 1 | 5 | 5 |  |
| 15. Visual Information | 26 | 3.93 (1.03) |  | 5 | 4 |  |
| 16. Credibility of Source | 35 | 4.50 (0.61) | 3 | 5 | 5 |  |

**Table S7.** Summary of open feedback of uMARS items.

| **uMars domain** | **Open feedback** |
| --- | --- |
| **Engagement** | Users liked the animation style of the app |
|  | Include more fun, engaging material |
|  | Include gamification features (e.g., word search exercise) |
|  | Include more customisation features (e.g., tree motif presented in different colours/backgrounds; more choice of background/text colour) |
|  | Diary consistently reported as the most helpful aspect of the app |
| **Functionality** | Navigation features of the app require improvement |
|  | Include speech-to-text functionality to overcome text-heavy nature of the app |
| **Aesthetics** | Liked the look-and-feel of the app metaphor and imagery (e.g. progress bars; tree motif flourishing with leaves as participant's worked through the app) |
|  | Add more customisable features |
| **Information** | The content of the app reflected users' difficulties and experiences |
|  | Inviting users to put themselves "in another's shoes" (i.e., to encourage mentalisation) helped users process personal experiences more objectively |
|  | Videos: soe users said the videos helpfully explained the content of the app; others felt the videos require improvement (e.g., clearer explanations, more accessible length) |
|  | Less "wordy"/text heavy |
|  | Include more trigger warnings in the app |

**Table S8.** Perceived impact of using the i-Minds app on mental health understanding.

|  |  | **Mean**<br>**(SD)** | **Min.** | **Max.** | **Median** | **Range of**<br>**possible**<br>**scores** |
| --- | --- | --- | --- | --- | --- | --- |
| *Using the i-Minds app...* |  |  |  |  |  |  |
| ... increased my awareness on the importance of my mental health | 36 | 3.40<br>(0.96) | 1 | 5 | 3 | 1-5 |
| .... increased my knowledge/understanding of my mental health | 36 | 3.51<br>(0.93) | 1 | 5 | 4 | 1-5 |
| ... has changed my attitudes towards improving my mental health | 36 | 3.13<br>(1.18) | 1 | 5 | 3 | 1-5 |
| ... has made me more motivated to address my mental health | 36 | 3.40<br>(1.12) | 1 | 5 | 4 | 1-5 |
| ... would encourage me to seek further help for my mental health (if I needed it) | 35 | 3.59<br>(1.12) | 1 | 5 | 4 | 1-5 |
| ... will make me do things/activities that will help my mental health | 36 | 3.21<br>(1.18) | 1 | 5 | 3 | 1-5 |

**Table S9.**
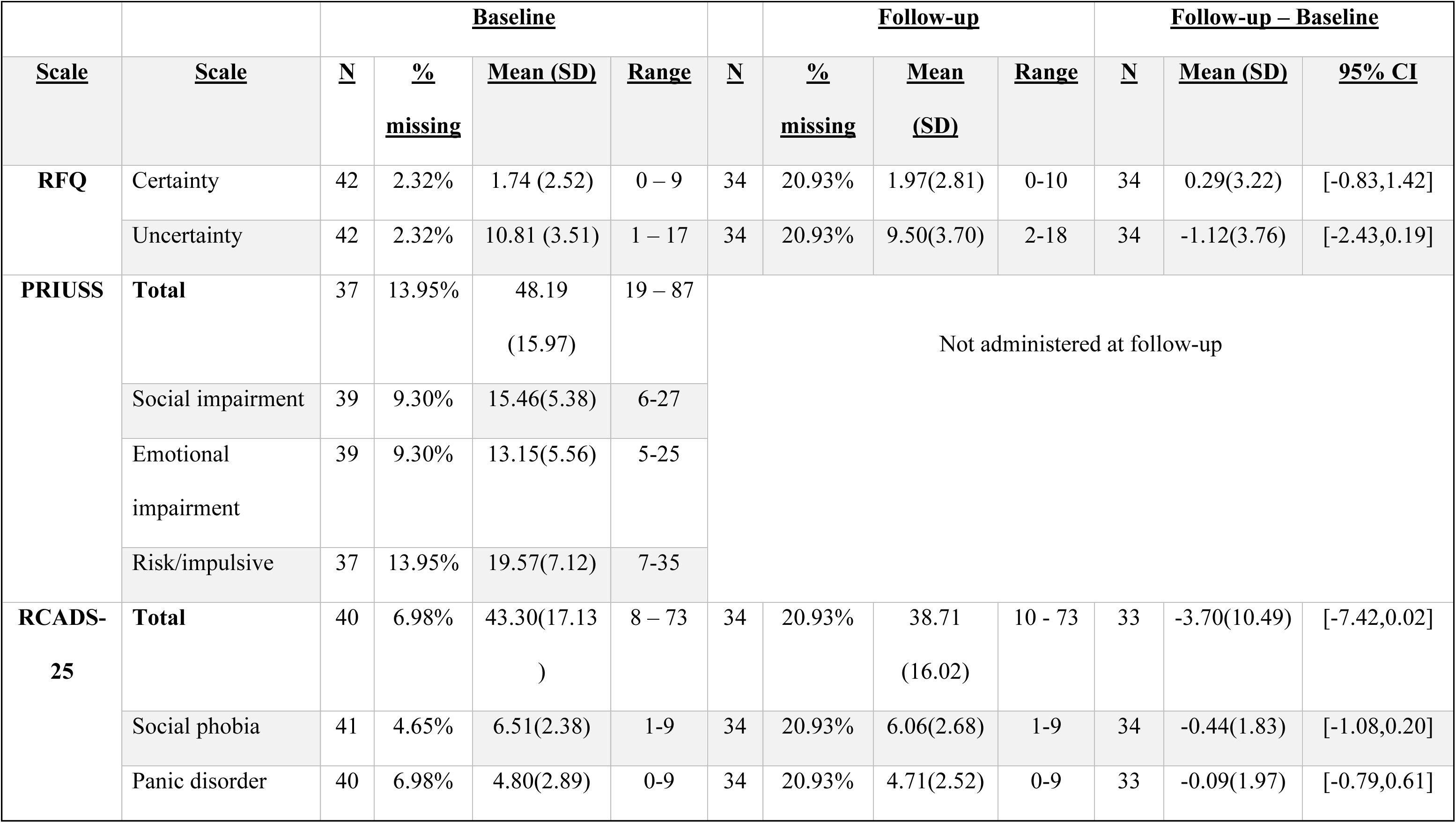
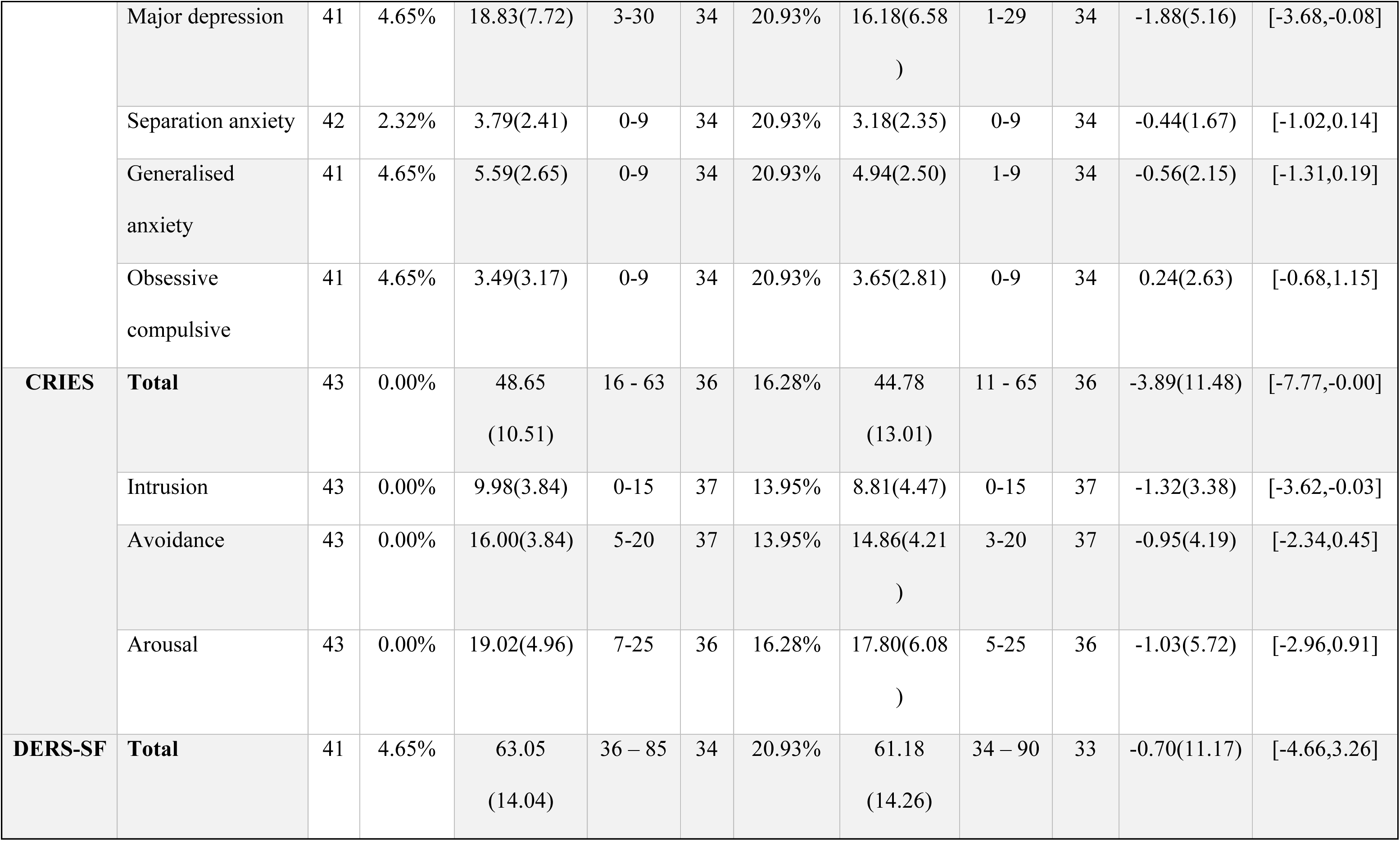
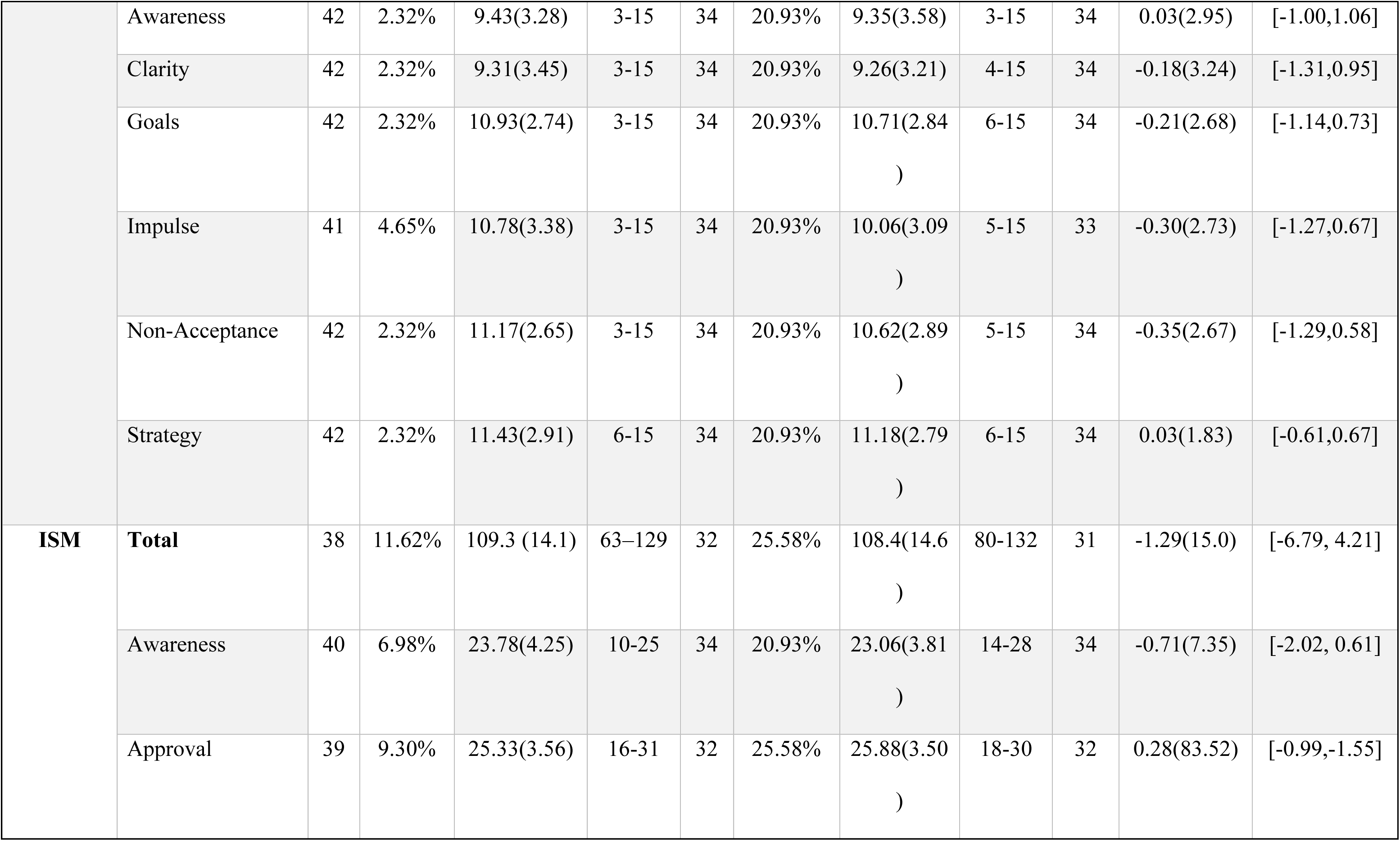
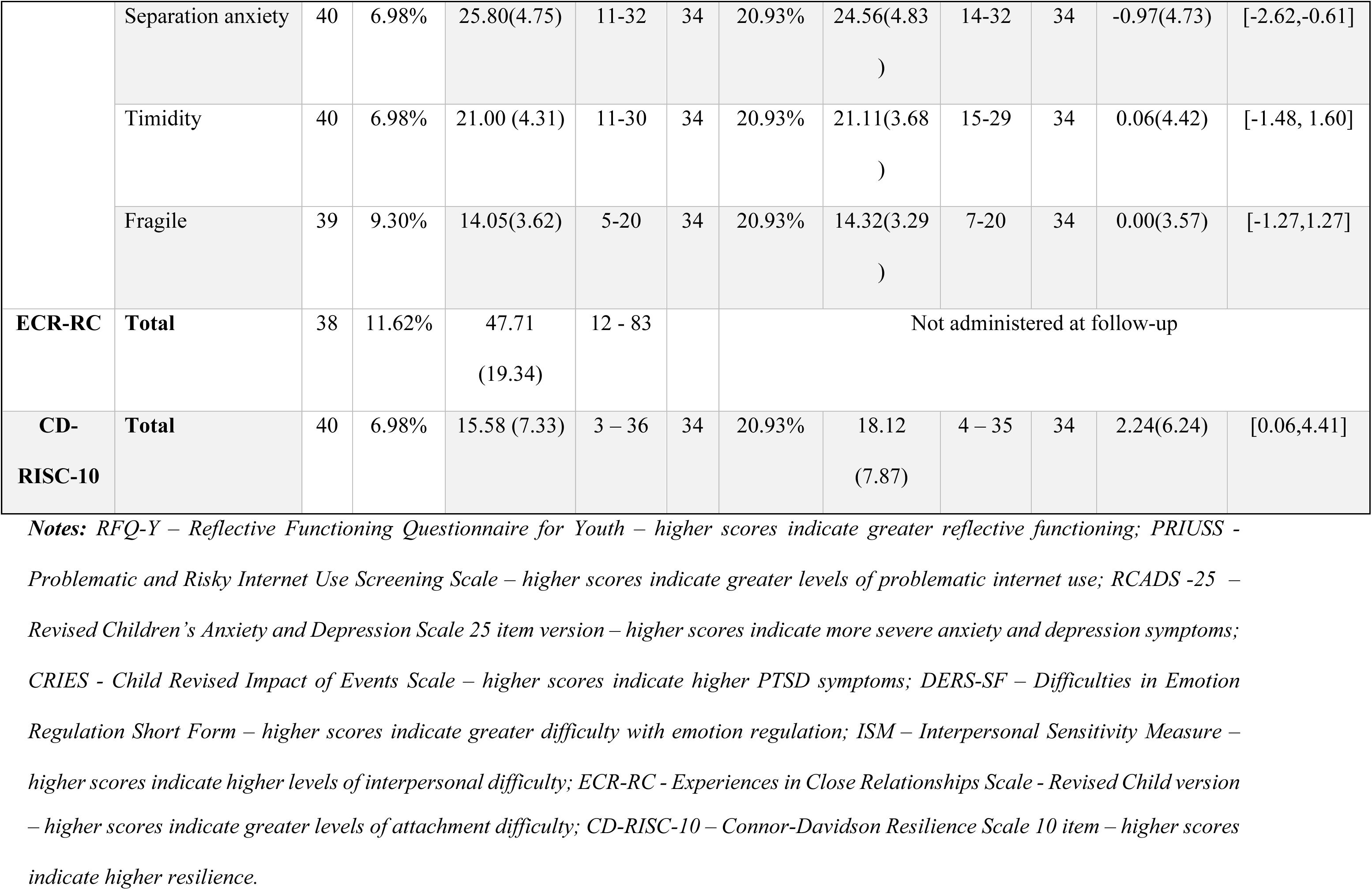
Mental health outcome data at baseline and follow-up assessments.

**Table S10.** Reliable change index analysis across clinical outcomes.

| Measure | Sample Size<br>(n) | No Change<br>n (%) | Improve<br>n (%) | Deteriorate<br>n (%) |
| --- | --- | --- | --- | --- |
| RFQ-C | 34 | 29 (85.3%) | 4 (11.8%) | 1 (2.9%) |
| RFQ-U | 34 | 20 (58.8%) | 10 (29.4%) | 4 (11.8%) |
| CRIES | 36 | 26 (72.2%) | 7 (19.4%) | 3 (8.3%) |
| DERS-SF | 33 | 24 (72.7%) | 5 (15.2%) | 4 (12.1%) |
| ISM | 31 | 21 (67.7%) | 5 (16.1%) | 5 (16.1%) |
| CD-RISC | 34 | 28 (82.4%) | 6 (17.6%) | 0 (0.0%) |

**Notes:** CRIES - Child Revised Impact of Events Scale; DERS-SF – Difficulties in Emotion Regulation Short Form; ISM – Interpersonal Sensitivity Measure; CD-RISC-10 – Connor-Davidson Resilience Scale 10 item; RFQ-C, Reflective Functioning Questionnaire for Youth-Certainty; RFQ-U, Reflective Functioning Questionnaire for Youth-Uncertainty.

